# Comparison Between Influenza and COVID-19 at a Tertiary Care Center

**DOI:** 10.1101/2020.08.19.20163857

**Authors:** Michael W. Donnino, Ari Moskowitz, Garrett S. Thompson, Stanley J. Heydrick, Rahul D Pawar, Katherine M. Berg, Shivani Mehta, Parth V. Patel, Anne V. Grossestreuer

**Author notes:** Corresponding Author: Michael W. Donnino, Center for Resuscitation Science, Beth Israel Deaconess Medical Center, 1 Deaconess Road, Rosenberg 2, Boston, MA 02215.

## Abstract

**Background:** Widespread reports suggest the characteristics and disease course of coronavirus disease 2019 (COVID-19) and influenza differ, yet detailed comparisons of their clinical manifestations are lacking.

**Objective:** Comparison of the epidemiology and clinical characteristics of COVID-19 patients with those of influenza patients in previous seasons at the same hospital

**Design:** Admission rates, clinical measurements, and clinical outcomes from confirmed COVID-19 cases between March 1 and April 30, 2020 were compared with those from confirmed influenza cases in the previous five influenza seasons (8 months each) beginning September 1, 2014.

**Setting:** Large tertiary care teaching hospital in Boston, Massachusetts

**Participants:** Laboratory-confirmed COVID-19 and influenza inpatients

**Measurements:** Patient demographics and medical history, mortality, incidence and duration of mechanical ventilation, incidences of vasopressor support and renal replacement therapy, hospital and intensive care admissions.

**Results:** Data was abstracted from medical records of 1052 influenza patients and 583 COVID-19 patients. An average of 210 hospital admissions for influenza occurred per 8-month season compared to 583 COVID-19 admissions over two months. The median weekly number of COVID-19 patients requiring mechanical ventilation was 17 (IQR: 4, 34) compared to a weekly median of 1 (IQR: 0, 2) influenza patient (p=0.001). COVID-19 patients were significantly more likely to require mechanical ventilation (31% vs 8%), and had significantly higher mortality (20% vs. 3%; p<0.001 for all). Relatively more COVID-19 patients on mechanical ventilation lacked pre-existing conditions compared with mechanically ventilated influenza patients (25% vs 4%, p<0.001).

**Limitation:** This is a single-center study which could limit generalization.

**Conclusion:** COVID-19 resulted in more hospitalizations, higher morbidity, and higher mortality than influenza at the same hospital.

## Introduction

COVID-19 and influenza are both highly infectious viral diseases characterized by pneumonia and acute respiratory failure in severe cases. Both place a burden on the health care system, but objective granular assessments of their comparative impact on individuals and the healthcare system are lacking in the literature.

The epidemiology, symptomology and annual public health burden presented by influenza is generally well characterized; however, this is largely through estimates as opposed to comprehensive reporting.^1^ The Centers for Disease Control and Prevention (CDC) estimates that over the past 10 years, between 9 and 45 million people have contracted influenza annually in the United States, with annual hospitalizations ranging from 140,000 – 810,000 and annual mortality ranging from 12,000 to 61,000 (between 0.10% and 0.17% of all cases), depending on the severity of the season.^2^ Influenza generally achieves community spread nationwide and can strike all age groups, but mortality is consistently higher in patients who are elderly, immunocompromised, and/or have pre-existing comorbidities such as chronic obstructive pulmonary disease, chronic kidney injury, cirrhosis, or cardiac disease.^3–9^ Decades of experience with influenza have led to vaccination programs which can mitigate the impact of influenza in years when the predominant circulating strain is accurately predicted.^10^

Official CDC counts indicate that over one million people in the United States have contracted COVID-19 between the virus’s first detected cases through April 30, 2020, with over 150,000 hospitalizations and over 63,000 deaths reported as of that date.^11^ However, the methods for reporting COVID-19 and influenza cases and outcomes differ in that COVID-19 is directly reported from each state whereas influenza cases are estimates based on reporting from selected hospitals.^1^ Moreover, granular comparisons between these two diseases such as rates and duration of respiratory failure (i.e., mechanical ventilation), need for hospitalization, outcomes of those patients without comorbid disease, frequency of non-pulmonary organ injury, and mortality are lacking, yet essential for understanding these two disease processes.

In this study, we compared the epidemiology and clinical characteristics of COVID-19 patients admitted to a large tertiary care teaching hospital in March and April of 2020 with those of influenza patients admitted to the same hospital over the prior five years.

## Methods

### Design

This was a single center, retrospective study at an urban tertiary care center. We compared patients admitted to the hospital with influenza during five influenza seasons (September-April 2014-2019) to patients admitted to the hospital with COVID-19 in March and April 2020. This study was reviewed by the Institutional Review Board at Beth Israel Deaconess Medical Center, which determined the study met exempt status. Written informed consent was therefore not required.

### Patient Population

Adult (aged >17 years) patients were included if they were admitted to the hospital for confirmed influenza or COVID-19 during the time frames of interest. Patients with influenza or COVID-19 were identified by ICD-10 codes (see appendix) and/or laboratory-based testing. Laboratory diagnosis of influenza and COVID-19 were carried out through specific real-time reverse transcriptase–polymerase chain reaction (RT-PCR) assays of nasal swab specimens. COVID-19 or influenza diagnosis for all subjects identified by ICD-10 codes were manually reviewed for confirmation. The ICD-10 codes are displayed in supplement Table S1.

### Study Data Abstraction

Study data was abstracted from patient electronic medical records based on the mechanism for identification of subjects described above. Data included demographics, personal medical histories (e.g. chronic comorbid conditions), laboratory findings on admission, intensive care unit admission status, intensive care unit length of stay, mechanical ventilation, hospital length of stay, and mortality. Key variables including presence and duration of mechanical ventilation and mortality were manually reviewed to verify the data. To compute volume of admissions between the two diseases, weekly counts of hospital admissions and mechanically ventilated patients were assessed during the peak two months of influenza seasons from 2014-2019 and from March-April 2020 for COVID-19. Reasons for intubation were manually categorized and all past medical history was manually extracted. All cases for which laboratory testing occurred greater than five days after admission were reviewed to determine if disease was believed to be nosocomial. For other variables (e.g., laboratory values), a sample of randomly chosen data was verified using manual chart review. Definitions for elements of the data abstraction are displayed in supplement Table S2.

### Data Analysis

Continuous data are presented as medians with interquartile ranges and categorical data are presented as counts and percentages. For continuous outcomes (duration of mechanical ventilation, duration of index hospital stay, number of weekly admissions, number of weekly incident mechanical ventilations), medians were compared using a Wilcoxon rank-sum test. For categorical outcomes (incidence of mechanical ventilation, incidence of shock, incidence of renal replacement therapy, hospital readmissions, ICU admissions, in-hospital mortality, reason for mechanical ventilation, and proportion of patients on mechanical ventilation with no major comorbidities), proportions were compared using chi-squared or Fisher’s exact tests, as appropriate. All analyses were done using Stata 14.2 (College Station, TX) and a p-value <0.05 was considered statistically significant.

## Results

### Overview of COVID-19 and Influenza Admissions with Key Outcomes

In total, 1855 patients were identified and 1635 were included in the study (Figure 1). Of these, 583 patients had laboratory-confirmed COVID-19 and 1052 patients had laboratory-confirmed influenza. An overview of admissions, intensive care unit admissions, receipt of mechanical ventilation, and mortality is illustrated per influenza season compared to COVID-19 (Table 1). The total admissions for influenza per eight-month influenza season averaged 210 compared to a total of 583 COVID-19 admissions over the two month COVID-19 study period. A total of 174 COVID-19 patients who were admitted in March and April 2020 were placed on mechanical ventilation compared to 84 patients admitted over five seasons of influenza. The proportion of admitted patients who received mechanical ventilation was significantly higher for COVID-19 patients compared to influenza patients (30% [174/583] versus 8% [84/1052], p< 0.001). One hundred and nineteen patients (20%) admitted in March and April 2020 with COVID-19 died compared to 34 patients (3%) over five seasons of influenza (p<0.001). Figure 2 illustrates the rates of COVID-19 admissions compared to influenza admissions per season in the busiest two months of each influenza season. The timing of local social distancing measures for COVID-19 is also indicated on Figure 2. There was a median of 70 (IQR: 13, 144) COVID-19 patients admitted per week compared to 16 (IQR: 8, 20) during the busiest two months of the influenza season (p=0.04). In addition, the number of COVID-19 patients placed on mechanical ventilation per week was 17 (IQR: 4, 34) compared to a median of 1 (IQR: 0, 2) influenza patient (p<0.001).

**Table 1.** Number of total patients, deaths, ICU admissions, and patients requiring mechanical ventilation in five influenza seasons (September 2014-May 2019) and the initial two months (March and April 2020) of COVID-19.

| Season | Total Patients<br>n | Mortality<br>n (%) | ICU admission<br>n (%) | Mechanical<br>Ventilation<br>n (%) |
| --- | --- | --- | --- | --- |
| Influenza (Season - September to April: 8 months) |  |  |  |  |
| 2014-2015 | 226 | 5 (2) | 54 (24) | 18 (8) |
| 2015-2016 | 134 | 3 (2) | 37 (28) | 15 (11) |
| 2016-2017 | 215 | 7 (3) | 34 (16) | 9 (4) |
| 2017-2018 | 268 | 10 (4) | 60 (22) | 25 (9) |
| 2018-2019 | 209 | 9 (4) | 46 (22) | 17 (8) |
| TOTALS | 1052 | 34 (3) | 231 (22) | 84 (8) |
| COVID-19 (Season - March to April: 2 months) |  |  |  |  |
| 2020 | 583 | 119 (20) | 251 (43) | 174 (30) |

p<0.001 for comparison between total influenza mortality and COVID-19 mortality, comparison between total influenza ICU admissions and COVID-19 admissions, and comparison between total influenza mechanical ventilations and COVID-19 ventilations.

**Figure 1.**
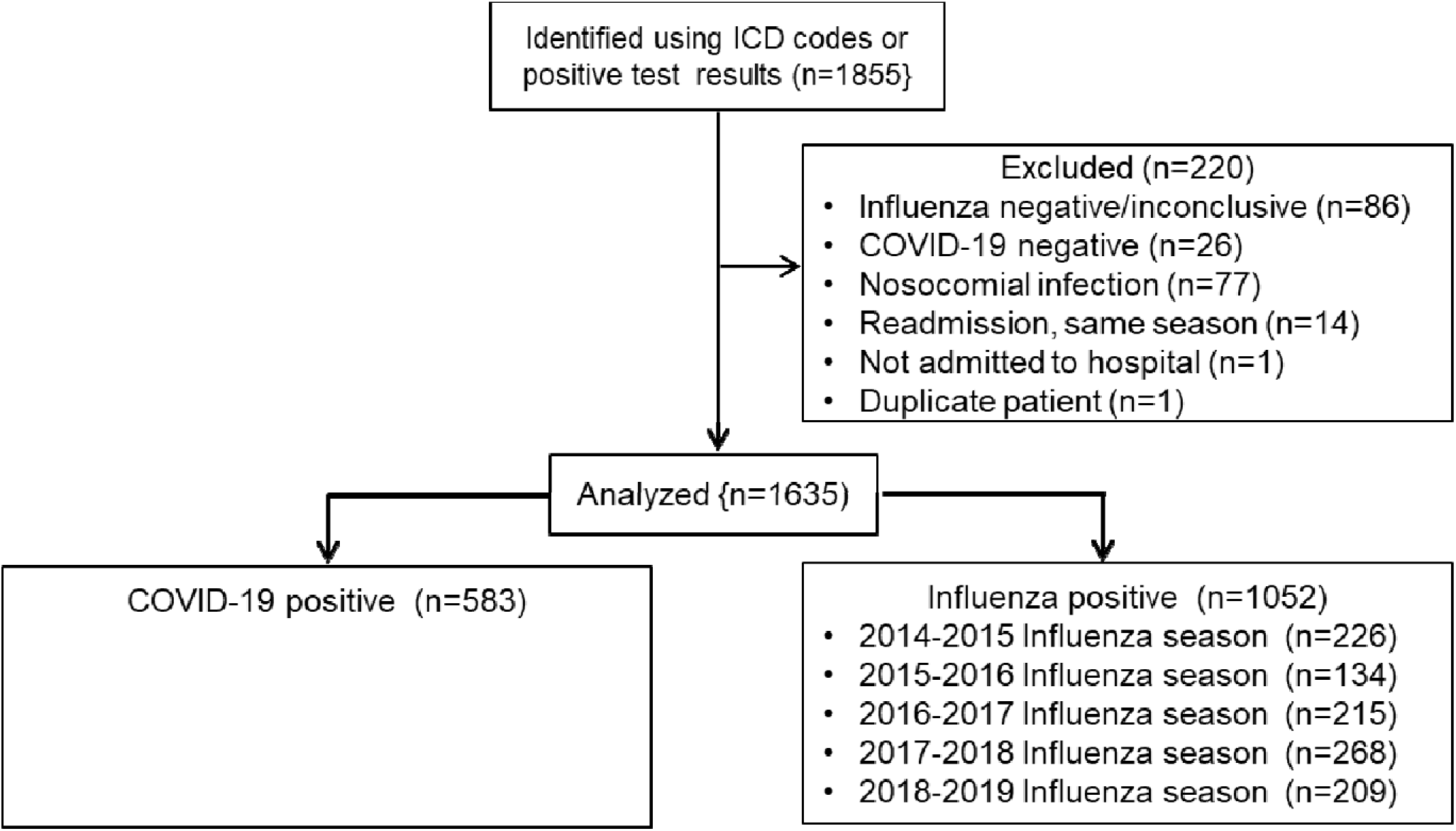
Patient Inclusion/Exclusion Criteria

**Figure 2.**
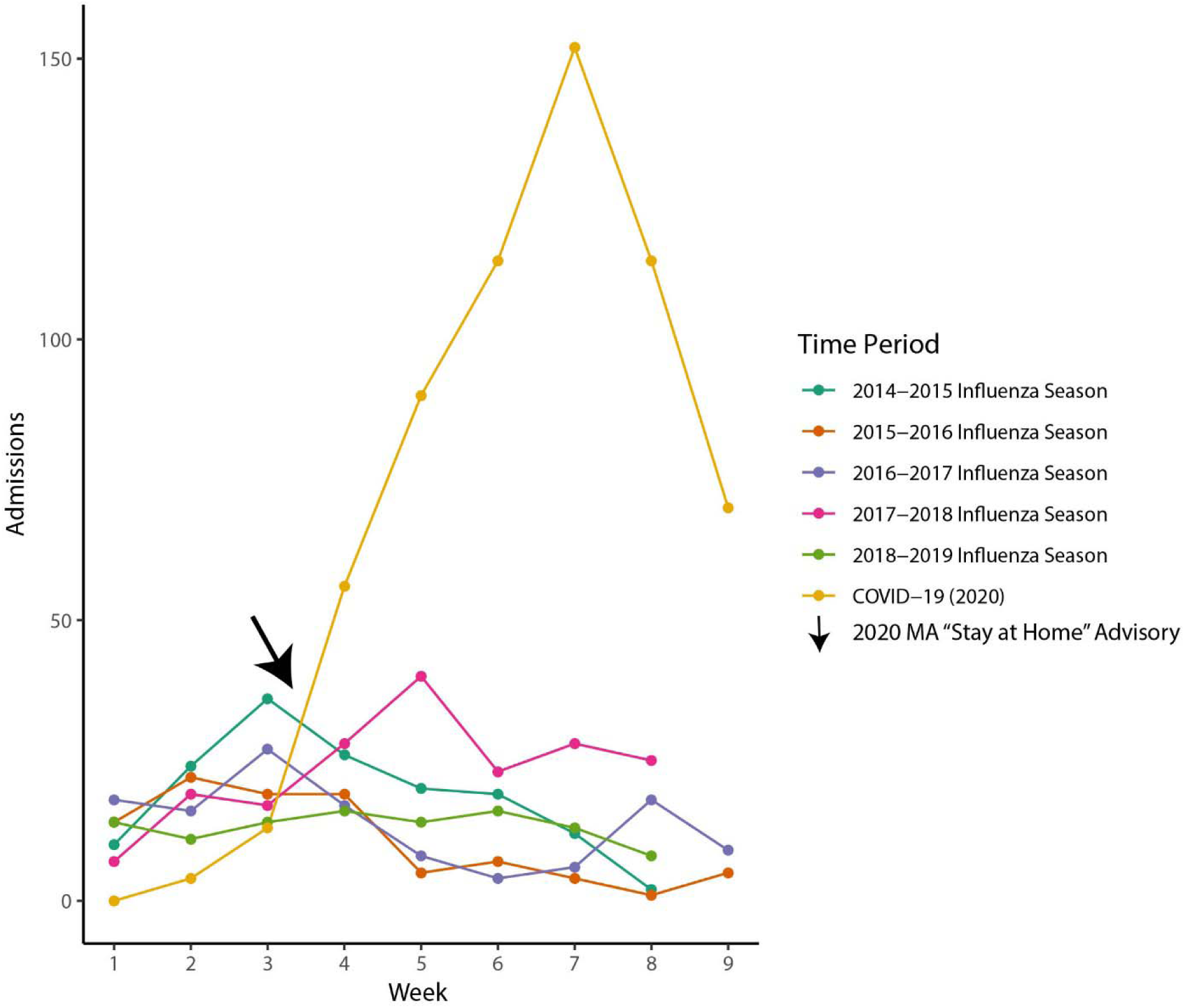
Comparison of COVID-19 hospital admissions during the 9 weeks in March-April 2020 and Influenza hospital admissions for the peak 9 weeks in five different seasons. The arrow denotes the approximate timing of the “Stay at Home” advisory put in place in Massachusetts on March 24, 2020.

### Demographics and Clinical Characteristics

Patient demographics and medical history are summarized in Table 2. Laboratory values for all patients and patients who required mechanical ventilation are displayed in supplement Tables S3 and S4. There were 304 (52%) males admitted with COVID-19 and 481 (46%) males admitted with influenza. The median age for COVID-19 admissions was 66 (IQR: 52, 77) and 68 (IQR: 56, 80) for influenza admissions. The median body mass index for COVID-19 patients was 29.3 (IQR: 25.7, 34.1) compared to 27.3 (IQR: 23.6, 32.3) for influenza. Past medical history is noted between the two groups in Table 2 for the COVID-19 cohort and the 2018-2019 season of influenza. There were significantly more patients with no major comorbidities in the COVID-19 group than in the influenza patients (113 [19%] vs 90 [9%], p<0.001).

**Table 2.** Demographics and Past Medical History

|  | **All patients** |  | **Mechanically ventilated patients** |  |
| --- | --- | --- | --- | --- |
|  | COVID-19<br>(n=583) | Seasonal<br>Influenza<br>(n=1052) | COVID-19<br>(n=174) | Seasonal<br>Influenza<br>(n=84) |
| **Demographics** |  |  |  |  |
| Male: n (%) | 304 (52) | 481 (46) | 96 (55) | 48 (57) |
| Age: median years (IQR) | 66 (52, 77) | 68 (56, 80) | 66 (54, 74) | 68 (57, 81) |
| Body Mass Index: median (IQR)* | 29.3<br>(25.7, 34.0) | 27.3<br>(23.6, 32.3) | 30.4<br>(27.3, 35.5) | 28.0<br>(23.3, 32.9) |
| **Past medical history** |  |  |  |  |
| No major comorbidities: n (%) | 113 (19) | 90 (9) | 44 (25) | 3 (4) |
| Cardiovascular disease: n (%) | 141 (24) | 373 (35) | 37 (21) | 27 (32) |
| Respiratory disease (COPD, asthma): n (%) | 119 (20) | 299 (28) | 29 (17) | 30 (36) |
| Dementia: n (%) | 69 (12) | 84 (8) | 10 (6) | 5 (6) |
| Stroke: n (%) | 50 (9) | 90 (9) | 8 (5) | 1 (1) |
| Immunosuppressed (Transplant, HIV/AIDS): n (%) | 10 (2) | 96 (9) | 3 (2) | 4 (5) |
| Hypertension: n (%) | 352 (60) | 638 (61) | 106 (61) | 48 (57) |
| Diabetes Mellitus: n (%) | 210 (36) | 298 (28) | 69 (39) | 30 (36) |
| Renal Disease: n (%) | 110 (19) | 211 (20) | 30 (17) | 15 (18) |
| Liver Disease: n (%) | 25 (4) | 73 (7) | 11 (6) | 1 (1) |
| Cancer: n (%) | 95 (16) | 279 (27) | 32 (18) | 24 (29) |
| Morbid Obesity (BMI>40) | 52 (11) | 77 (8) | 23 (14) | 4 (6) |
| Alcohol Use Disorder: n (%) | 33 (6) | 147 (14) | 16 (9) | 7 (8) |
| Tobacco: n (%) | 107 (18) | 211 (20) | 29 (17) | 22 (26) |

\*BMIs were missing from 77 patients with COVID-19 and 22 patients with influenza

In patients who received mechanical ventilation, 55% of the COVID-19 patients were male, compared to 57% of the influenza patients. The median age for COVID-19 patients on mechanical ventilation was 66 (IQR: 54, 74), compared with 68 (IQR: 57, 81) for influenza patients on mechanical ventilation. The median body mass index for COVID-19 patients on mechanical ventilation was 30.8 (IQR: 27.3, 36.1) compared to 27.6 (IQR: 23.3, 32.6) for influenza patients. COVID-19 patients who were mechanically ventilated were statistically significantly more likely to not have major comorbidities compared to influenza patients on mechanical ventilation (44 [25%] vs 3 [4%], p<0.001). The reasons for intubation and subsequent mechanical ventilation are displayed in Table 3. Overall, the reasons for intubation in COVID-19 patients were different than those intubated for influenza (p<0.001). Pneumonia and/or acute respiratory distress syndrome (ARDS) was the reason for 94% of intubations in COVID-19 patients, while this accounted for only 56% of intubations in influenza patients. Additionally, no COVID-19 patients received mechanical ventilation due to exacerbation of pre-existing conditions compared to 14% of influenza cases who received mechanical ventilation for this indication.

**Table 3.** Reasons for respiratory failure in patients with COVID-19 in March-April 2020 compared to patients with influenza in five influenza seasons (September 2014-May 2019)

|  | COVID-19<br>(n=175) | Influenza<br>(n=84) | p-value |
| --- | --- | --- | --- |
| Pneumonia /Acute Respiratory Distress Syndrome: n (%) | 164 (94) | 47 (56) | <0.001 |
| Exacerbation of pre-existing health condition<br>(e.g., asthma, chronic lung disease, etc.): n (%) | 0 (0) | 12 (14) |  |
| <u>Other Causes:</u> n (%) | 10 (6) | 17 (30) |  |
| Neurological (e.g., intracranial bleed) | 1 (1) | 4 (5) |  |
| Surgery/procedure | 2 (1) | 8 (10) |  |
| Cardiac arrest | 2 (1) | 1 (1) |  |
| Gastrointestinal hemorrhage | 1 (1) | 2 (2) |  |
| Hemodynamic instability | 0 (0) | 1 (1) |  |
| Trauma | 0 (0) | 1 (1) |  |
| Unknown | 1 (1) | 0 (0) |  |
| Multifactorial causes not deemed to be primarily pneumonia | 3 (2) | 8 (10) |  |

### Additional Outcomes

Table 4 summarizes additional clinical outcomes including duration of index hospital admission, duration of mechanical ventilation, provision of vasopressor support, and provision of renal replacement therapy. Patients with COVID-19 had a statistically significantly longer median duration of mechanical ventilation than patients with influenza (14.3 [IQR: 7.9, 22.5] vs 3.3 [IQR: 1.7, 7.9] days, p<0.001). Fifty COVID-19 patients (9%) received renal replacement therapy during their hospitalization, compared to 12 (1%) of influenza patients (p<0.001). One hundred and seventy-seven COVID-19 patients (29%) received vasopressor support during their hospitalization, compared to 74 (7%) of influenza patients (p<0.001). Six of the COVID-19 deaths during the 2 month COVID-19 window were patients who had no major comorbidities (1% of total patients), whereas potentially only three deaths during five years of influenza seasons were patients without comorbidities (0.3% of total patients). No deaths among those without major comorbidities were reported in the most recent influenza season. Of note, one of the three deaths in the influenza cohort occurred in a patient who presented with and died from widespread arterial and venous thrombus of unclear etiology and had not seen a physician in many years thus making a determination of underlying conditions difficult but was given a default labeling of healthy.

**Table 4.** Clinical outcomes in COVID-19 patients treated between March-April 2020 and influenza patients treated during five influenza seasons (September 2014-May 2019)

| **Outcome** | **COVID-19**<br>**Spring, 2020**<br>**( n= 583)** | **Influenza**<br>**2014-2019**<br>**(n =1052)** | **p-value** |
| --- | --- | --- | --- |
| Index hospital length of stay: median days (IQR) | 8 (4, 16) | 4 (2, 7) | <0.001 |
| Mechanical ventilation: n (%) | 174 (30) | 84 (8) | <0.001 |
| Time on mechanical ventilation - intubated patients:*<br>median days (IQR) | 14.3 (7.9, 22.5) | 3.3 (1.7, 7.9) | <0.001 |
| Time on mechanical ventilation - intubated patients with<br>pneumonia/ARDS:* median days (IQR) | 14.7 (8.5, 22.7) | 3.8 (2.0, 12.5) | <0.001 |
| Shock - on vasopressors during hospitalization**: n (%) | 177 (29) | 74 (7) | <0.001 |
| Renal replacement therapy: n (%) | 50 (9) | 12 (1) | <0.001 |
| Readmitted within 30 days: n (%) | 29 (5) | 15 (1) | <0.001 |
| Hospital mortality: n (%) | 119 (20) | 34 (3) | <0.001 |

\*including reintubation time
\*\*measured at the encounter level (n=613 for COVID-19; n=1070 for influenza)

## Discussion

COVID-19 has been compared to influenza by both health care professionals and the lay public ^12–16^ but limited detailed objective data are available for comparing and contrasting the impact of these two disease processes on patients and hospitals. We found that admissions for COVID-19 over a two month period at our medical center were more than double the total number of admissions for influenza during any 8-month influenza season in the past five years. In addition to the larger volume of cases within a much shorter time period, severity of illness and lethality for COVID-19 were also markedly higher than for influenza. We observed more mechanically ventilated COVID-19 patients (i.e., those with severe, life-threatening illness) in a two month period than occurred in five entire seasons of influenza combined, and a similar observation was made with patient deaths. Taken together, these findings indicate that COVID-19 causes more severe disease and is more lethal than influenza. Other studies aimed at total incidence (hospitalized plus non-hospitalized) suggest a higher rate of spread (Ro) of COVID-19 than influenza^17–19^ and our markedly increased hospital admissions are consistent with this.

COVID-19 resulted in not only a marked increase in the number of mechanically ventilated patients in a short period of time but also a longer median duration of mechanical ventilation within this population. Specifically, 174 patients required mechanical ventilation for COVID-19 with a median duration on the ventilator of 14 days. In contrast, the combined seasons of influenza only resulted in 84 mechanically ventilated patients for a median duration of 3 days. This combination illustrates both the acute severity of disease and the prolonged nature of the respiratory failure that occurs in COVID-19 compared to influenza.

COVID-19 caused a substantial number of patients without major comorbid disease to require mechanical ventilation (44 people in 2 months). In contrast, influenza rarely led to mechanical ventilation in patients without underlying comorbidities (3 people over five seasons). In this specific subgroup, five patients with COVID-19 died in the two-month period compared to one with influenza over five years. Pneumonia and acute respiratory distress syndrome were the predominant causes of mechanical ventilation for COVID-19 (94%), whereas this was not the case for influenza (55%). With influenza, the need for mechanical ventilation often developed as a result of exacerbation of a pre-existing health condition such as asthma or chronic obstructive pulmonary disease (COPD), which did not occur in COVID-19. The apparently higher rate of pneumonia and ARDS in our COVID-19 cohort may at least partly explain the more frequent occurrence of respiratory failure and death even in the absence of serious comorbidities.

In addition to acute respiratory failure, rates of vasopressor and renal replacement therapy were significantly increased in COVID-19 compared to influenza. The increased disease severity reflected in the rates of acute respiratory failure and other-organ injury paralleled the overall higher lethality of COVID-19 which resulted in 119 deaths in 2 months compared to 34 deaths from influenza over five seasons. The disparity in deaths was driven by both increased volume of cases and increased lethality within that volume in COVID-19 patients. This study did not evaluate organ-injury such as liver failure, neurological injury, and coagulation disorders, all of which have been reported with COVID-19.^20–26^

The increase in volume of COVID-19 cases compared to influenza is noted in Figure 2. This increase in patient volume occurred despite the implementation of increasingly stringent social distancing in Boston starting on March 15, 2020^27^ and a statewide stay-at-home advisory starting on March 24, 2020.^28^ New cases began to decrease approximately 6 weeks after these measures were taken. Thus, the overall volume of cases of COVID-19 in this report was likely modified by these measures, whereas these mitigation measures were not taken during any influenza season. Influenza, however, can be modified by the implementation of widespread vaccination programs which are currently not available for COVID-19.

While not a focus of this study, the critical care resources required to manage the marked increase and severity of disease in COVID19 included the emergent conversion of surgical and cardiac intensive care units to medical intensive care units, and the conversion of post-anesthesia care units and multiple medical wards to intensive care units. The conversion of any such space to an intensive care unit has not been done for any influenza season. In addition to space conversion, human resources (i.e., staffing models for new intensive care units), personal protective equipment, and increased use of dialysis were all part of the hospital-wide response to the COVID-19 pandemic but we did not compare these to influenza. Future studies may benefit from direct comparisons of resource utilization for the two disease processes.

Our study was limited by evaluation of subjects in one hospital system, though this methodology allowed for granular assessments of differences between diseases. In addition, the true impact of COVID-19 in the absence of social distancing measures remains unclear as these measures were taken early and within weeks of the onset of hospital admissions.

## Conclusions

COVID-19 resulted in significantly more weekly hospital admissions over two months than in five seasons of influenza in a region with community spread of both diseases. In addition, COVID-19 resulted in a significant increase in use of mechanical ventilation, an increase in the use of vasopressor support, an increase in incidence of renal injury, and an increase in mortality. COVID-19 was also significantly more likely to result in mechanical ventilation for those without major comorbidities as compared to influenza.

## Data Availability

THe data is maintained by the authors who can be contacted for further queries.

## Acknowledgements

The authors would like to thank Lethu A. Ntshinga, AB, Jacob Boise, BS, Emma Hershey, BA, Mahmoud S. Issa, MD, Ying K. Loo, BS and Natia Peradze, MD, PhD, for their data abstraction and data entry.

